# A comparison of five epidemiological models for transmission of SARS-CoV-2 in India

**DOI:** 10.1101/2020.09.19.20198010

**Authors:** Soumik Purkayastha, Rupam Bhattacharyya, Ritwik Bhaduri, Ritoban Kundu, Xuelin Gu, Maxwell Salvatore, Swapnil Mishra, Bhramar Mukherjee

## Abstract

Many popular disease transmission models have helped nations respond to the COVID-19 pandemic by informing decisions about pandemic planning, resource allocation, implementation of social distancing measures and other non-pharmaceutical interventions. We study how five epidemiological models forecast and assess the course of the pandemic in India: a baseline model, an extended SIR (eSIR) model, two extended SEIR (SAPHIRE and SEIR-*fansy*) models, and a semi-mechanistic Bayesian hierarchical model (ICM). Using COVID-19 data for India from March 15 to June 18 to train the models, we generate predictions from each of the five models from June 19 to July 18. To compare prediction accuracy with respect to reported cumulative and active case counts and cumulative death counts, we compute the symmetric mean absolute prediction error (SMAPE) for each of the five models. For active case counts, SMAPE values are 0.72 (SEIR-fansy) and 33.83 (eSIR). For cumulative case counts, SMAPE values are 1.76 (baseline) 23.10 (eSIR), 2.07 (SAPHIRE) and 3.20 (SEIR-*fansy*). For cumulative death counts, the SMAPE values are 7.13 (SEIR-*fansy*) and 26.30 (eSIR). For cumulative cases and deaths, we compute Pearson’s and Lin’s correlation coefficients to investigate how well the projected and observed reported COVID-counts agree. Three models (SAPHIRE, SEIR-*fansy* and ICM) return total (sum of reported and unreported) counts as well. We compute underreporting factors as of June 30 and note that the SEIR-*fansy* model reports the highest underreporting factor for active cases (6.10) and cumulative deaths (3.62), while the SAPHIRE model reports the highest underreporting factor for cumulative cases (27.79).

## 1. INTRODUCTION

Coronavirus disease 2019 (COVID-19) is an infectious disease caused by severe acute respiratory syndrome coronavirus 2 (SARS-CoV-2) (1). At the time of writing this paper, it has been identified as an ongoing pandemic, with more than 18 million reported cases across 188 countries and territories. The disease was first identified in Wuhan, Hubei, China in December 2019 (2). Since then, more than 936,000 lives have been lost and 20.1 million recoveries have been reported as a direct consequence of the disease. Notable outbreaks were recorded in the United States of America, Spain, Italy, Iran, Brazil and India -- which was a crucial battleground against the outbreak. The Indian government imposed very strict lockdown measures in order to attenuate the spread of the virus. Said measures have not been as effective as was intended (3), with India now reporting the largest number of confirmed cases in Asia, and the third highest number of confirmed cases in the world after the United States and Brazil (4), with the number of confirmed cases crossing the 1 million mark on July 17, 2020. On March 24, the Government of India ordered a 21-day nationwide lockdown, later extending it till May 3. This was followed by two-week extensions starting May 3 and 17 with substantial relaxations. From June 1, the government started ‘unlocking’ most regions of the country in three unlock phases. In order to formulate and implement policy geared toward containment and mitigation, it is important to recognize the presence of highly variable contagion patterns across different Indian states (5). There is a rising interest in studying potential trajectories that the infection can take in India to improve policy decisions.

A spectrum of models for projecting infectious disease spread have become widely popular in wake of the pandemic. Some popular models include the ones developed at the Institute of Health Metrics (IHME) (6) (University of Washington, Seattle) and at the Imperial College London (7). The IHME COVID-19 project initially relied on an extendable nonlinear mixed effects model for fitting parametrized curves to COVID-data, before moving to a compartmental model to analyze the pandemic and generate projections. The Imperial College model (henceforth ICM) works backwards from observed death counts to estimate transmission that occurred several weeks ago, allowing for the time lag between infection and death. A Bayesian mechanistic model is introduced - linking the infection cycle to observed deaths, inferring the total population infected (attack rates) as well as the time-varying reproduction number *R*(*t*). With the onset of the pandemic, there has been renewed interest in multi-compartment models, which have played a central role in modeling infectious disease dynamics since the 20^th^ century (8). The simplest of compartmental models include the standard SIR (9) model, which has been extended (10) to incorporate various types of time-varying quarantine protocols, including government-level macro isolation policies and community-level micro inspection measures. Further extensions include one which adds a spatial component to this temporal model by making use of a cellular automata structure (11). Larger compartmental models include those which incorporate different states of transition between susceptible, exposed, infected and removed (SEIR) compartments, which have been used in the early days of the pandemic in the Wuhan province of China (12). The SEIR compartmental model has been further extended to the SAPHIRE model (13), which accounts for the infectiousness of asymptomatic (14) and pre-symptomatic (15) individuals in the population (both of which are crucial transmission features of COVID-19), time varying ascertainment rates, transmission rates and population movement.

Researchers and policymakers are relying on these models to plan and implement public health policies at the national and local levels. New models are emerging rapidly. Models often have conflicting messages and it is hard to distinguish a good model from an unreliable one. Different models operate under different assumptions and provide different deliverables. In light of this, it is important to investigate and compare the findings of various models on a given test dataset. While some work has been done in terms of trying to reconcile results from different models of disease transmission that can be fit to data emerging from local and national governments (16), more comparisons need to be done to investigate how differences between competing models might lead to differing projections. In the context of India, such head-to-head comparison across models are largely unavailable.

We consider five different models of different genre, starting from the simplest baseline model. The baseline model we investigate relies on curve-fitting methods, with cumulative number of infected cases modeled as exponential process. Next, we consider the extended SIR (eSIR) model (10), which uses a Bayesian hierarchical model to generate projections of proportions of infected and removed people at future time points. The SAPHIRE (13) model has been demonstrated to reconstruct the full-spectrum dynamics of COVID-19 in Wuhan between January and March 2020 across 5 periods defined by events and interventions. Using the same model, we study the evolution of the pandemic in India over 4 well-defined lockdown periods, each with distinct transmission and ascertainment features. Another model (henceforth, SEIR-*fansy*) modifies the SEIR model to account for high false negative rate and symptom-based administration of COVID-19 tests. Finally, we study the ICM model, which utilizes a semi-mechanistic Bayesian hierarchical model based on renewal equations that model infections as a latent process and links deaths to infections with help of survival analysis. Each of the models mentioned above have had appreciable success in being able to satisfactorily analyze and project the trajectory of the pandemic in different countries (17),(18),(19).

In order to fairly compare and contrast the models mentioned above, we study their respective treatment of the different lockdown periods imposed by the Government of India. Additionally, we compare their projections based on reported data, with special emphasis on how the models deal with (if they do, at all) under-reporting and under-detection of COVID-cases, which has been a major point of discussion in the scientific community (20).

The rest of the paper is organized as follows. In *Section 2* we provide an overview of the various models considered in our analysis. The supplement has detailed discussion on the formulation, assumptions and estimation methods utilized by each of the models. We present the findings of our comparative investigation of the models in *Section 3* and discuss the implications of our findings in *Section 4*.

## 2. METHODS

### 2.1. Overview of models

In this section, we discuss the assumptions and formulation of each of the five models described above.

#### 2.1.a. Baseline model

#### Overview

The baseline model we investigate aims to predict the evolution of the COVID-19 pandemic by means of a regression-based predictive model (21). More specifically, the model relies on a regression analysis of the daily cumulative count of infected cases based on the least-squares fitting. In particular, the growth rate of the infection is modeled as an exponentially decaying process. *Figure 1* provides a schematic overview of this model.

**Figure 1:**
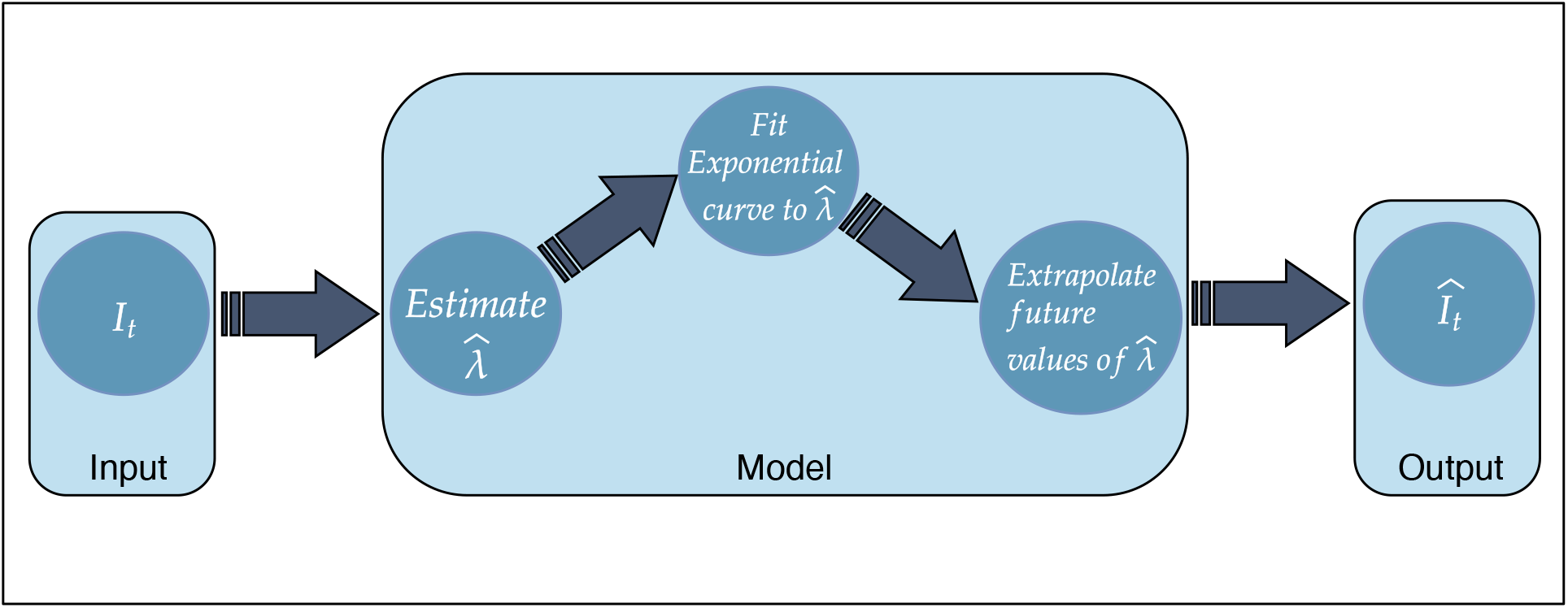
Schematic overview of the baseline model.

#### Formulation

The baseline model assumes that the following differential equation governs the evolution of a disease in a fixed population

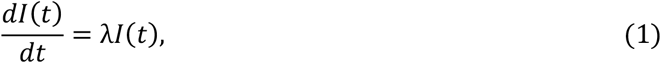

where *I*(*t*) is defined as the number of infected people at time *t* and λ is the growth rate of infection. Unlike the other models described in subsequent sections, the baseline model analyses and projects only the cumulative number of infections, and not counts/proportions associated with other compartments. The model uses reported field data of the infections in India over a specific time period. The growth rate can be numerically approximated from Equation (1) above as

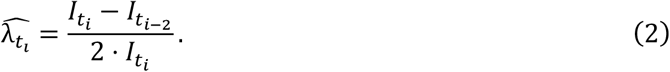

Having estimated the growth rate, the model uses a least-squares method to fit an exponential time-varying curve to 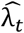, obtained from Equation (2) above. Using projected values of 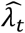, we extrapolate the number of infections which will occur in future.

#### Implementation

The baseline model described above has been implemented in R using standard packages.

#### 2.1.b. Extended SIR (eSIR) model

##### Overview

We use an extension of the standard susceptible-infected-removed (SIR) compartmental model known as the extended SIR (eSIR) model (10). To implement the eSIR model, a Bayesian hierarchical framework is used to model time series data on the proportion of individuals in the infected and removed compartments. Markov chain Monte Carlo (MCMC) methods are used to implement this model, which provides not only posterior estimation of parameters and prevalence values associated with all three compartments of the SIR model, but also predicted proportions of the infected and the removed people at future time points. *Figure 2* is a diagrammatic representation of the eSIR model.

**Figure 2:**
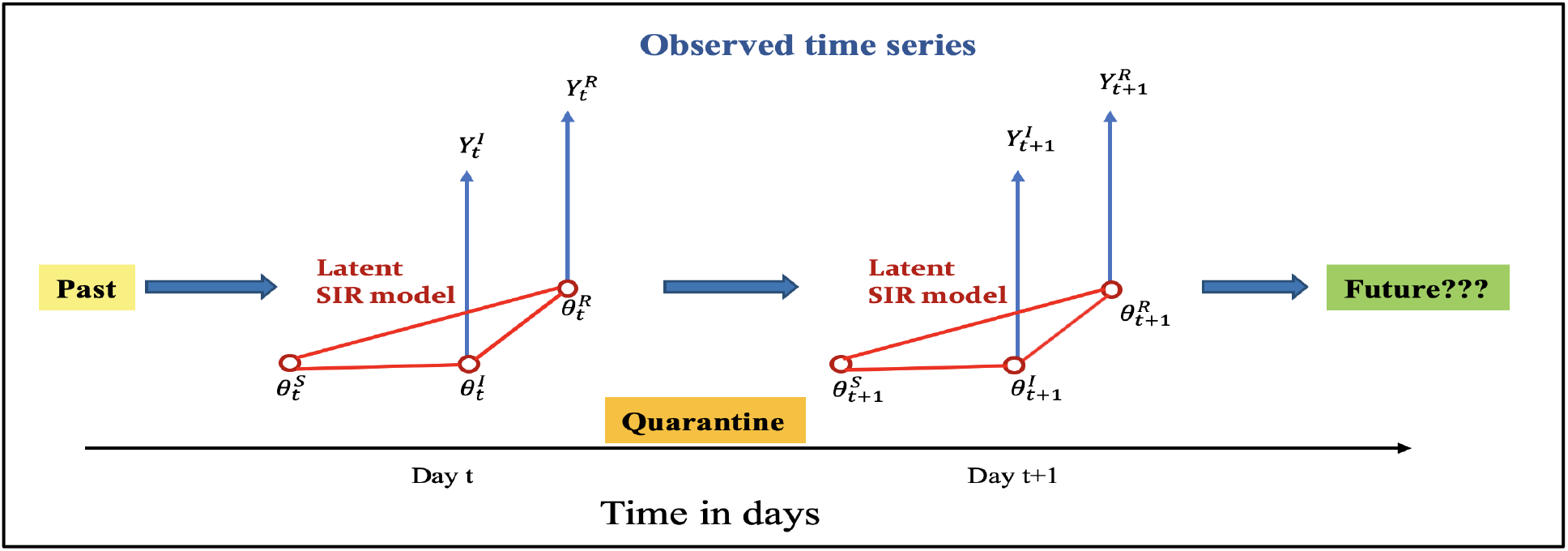
The eSIR model with a latent SIR model on the unobserved proportions. Reproduced from Wang et al., 2020 (10).

##### Formulation

The eSIR model assumes the true underlying probabilities of the three compartments follow a latent Markov transition process and require observed daily proportions of infected and removed cases as input.

The observed proportions of infected and removed cases on day *t* are denoted by 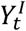 and 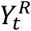, respectively. Further, we denote the true underlying probabilities of the S, I, and R compartments on day *t* by 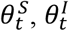 and 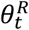, respectively, and assume that for any *t*,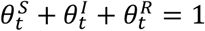. Assuming a usual SIR model on the true proportions we have the following set of differential equations

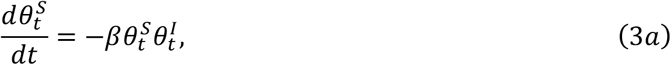

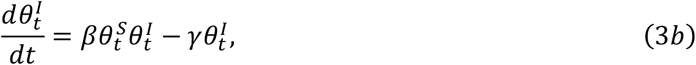

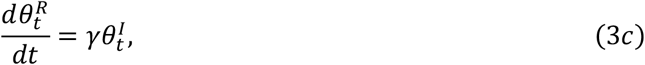

where *β* > 0 denotes the disease transmission rate, and *γ* > 0 denotes the removal rate. The basic reproduction number *R*_0_ := *β*/*γ* indicates the expected number of cases generated by one infected case in the absence of any intervention and assuming that the whole population is susceptible. We assume a Beta-Dirichlet state space model for the observed infected and removed proportions, which are conditionally independently distributed as

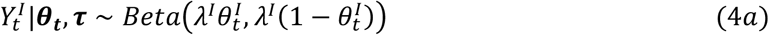

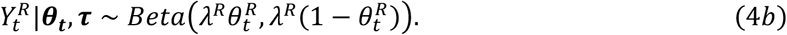

Further, the Markov process associated with the latent proportions is built as:

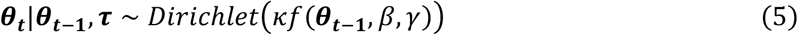

where ***θ***_***t***_ denotes the vector of the underlying population probabilities of the three compartments, whose mean is modeled as an unknown function of the probability vector from the previous time point, along with the transition parameters. 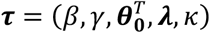 denotes the whole set of parameters where *λ*^*I*^, *λ*^*R*^ and *k* are parameters controlling variability of the observation and latent process, respectively. The function *f*(·) is then solved as the mean transition probability determined by the SIR dynamic system, using a fourth order Runge-Kutta (RK4) approximation (22).

##### Priors and MCMC algorithm

The prior on the initial vector of latent probabilities is set as 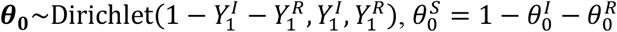. The prior distribution of the basic reproduction number is lognormal such that *E*(*R*_0_) = 3.28 (23). The prior distribution of the removal rate is also lognormal such that *E*(*γ*) = 0.5436. We use the proportion of death within the removed compartment as 0.0184 so that the infection fatality ratio is 0.01 (24). For the variability parameters, the default choice is to set large variances in both observed and latent processes, which may be adjusted over the course of epidemic with more data becoming available: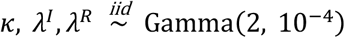.

Denoting *t*_0_ as the last date of data availability, and assuming that the forecast spans over the period [*t*_0_ + 1, *T*], our algorithm is as follows.

Step 0. Take *M* draws from the posterior 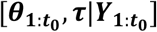.

Step 1. For each solution path *m* ∈ {1, …, *M*}, iterate between the following two steps via MCMC.

i. Draw 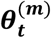 from 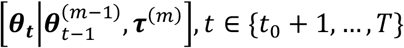.

ii. Draw 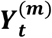 from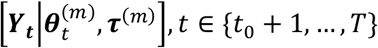.

##### Implementation

We implement the proposed algorithm in R package *rjags* (25) and the differential equations were solved via the fourth-order Runge–Kutta approximation. To ensure the quality of the MCMC procedure, we fix the adaptation number at 10^4^, thin the chain by keeping one draw from every 10 random draws to reduce autocorrelation, set a burn-in period of 10^5^ draws to let the chain stabilize, and start from 4 separate chains. Thus, in total, we have 2 × 10^5^ effective draws with about 2 × 10^6^ draws discarded. This implementation provides not only posterior estimation of parameters and prevalence of all the three compartments in the SIR model, but also predicts proportions of the infected and the removed people at future time point(s). The R package for implementing this general model for understanding disease dynamics is publicly available at *https://github.com/lilywang1988/eSIR*.

#### 2.1.c. SAPHIRE model

##### Overview

This model (13) extends the classic SEIR model to estimate COVID-related transmission parameters, in addition to projecting COVID-19 counts, while accounting for pre-symptomatic infectiousness, time-varying ascertainment rates, transmission rates and population movements. *Figure 3* provides a schematic diagram of the compartments and transitions conceptualized in this model. The model includes seven compartments: susceptible (S), exposed (E), pre-symptomatic infectious (P), ascertained infectious (I), unascertained infectious (A), isolation in hospital (H) and removed (R). Compared with the classic SEIR model, SAPHIRE explicitly models population movement and introduce two additional compartments (A and H) to account for the fact that only ascertained cases would seek medical care and thus be quarantined by hospitalization. The model described and implemented here relies on the same methodology and arguments as presented by the authors of the SAPHIRE model. The only difference is that while the original model analyzed data from China over a time period of December 2019 to March 2020 (which constituted the initial days of the pandemic in China), we analyze data from India, for the initial period of the pandemic in India. Additionally, the original manuscript adjusted the model to account for population movement. With the lockdown in India being severe (several states closed their respective borders) and data on population movement not being available, we make no such modifications. Additionally, described in greater detail in the subsequent sections, we note that the SAPHIRE model returns reported and unreported cumulative COVID-case counts, in addition to cumulative counts of the removed compartment. As such, for the purpose of comparisons, the SAPHIRE model is used only to study cumulative COVID-case counts, and not active case counts or cumulative death counts. The R package for implementing this general model for understanding disease dynamics is publicly available at *https://github.com/chaolongwang/SAPHIRE*.

**Figure 3:**
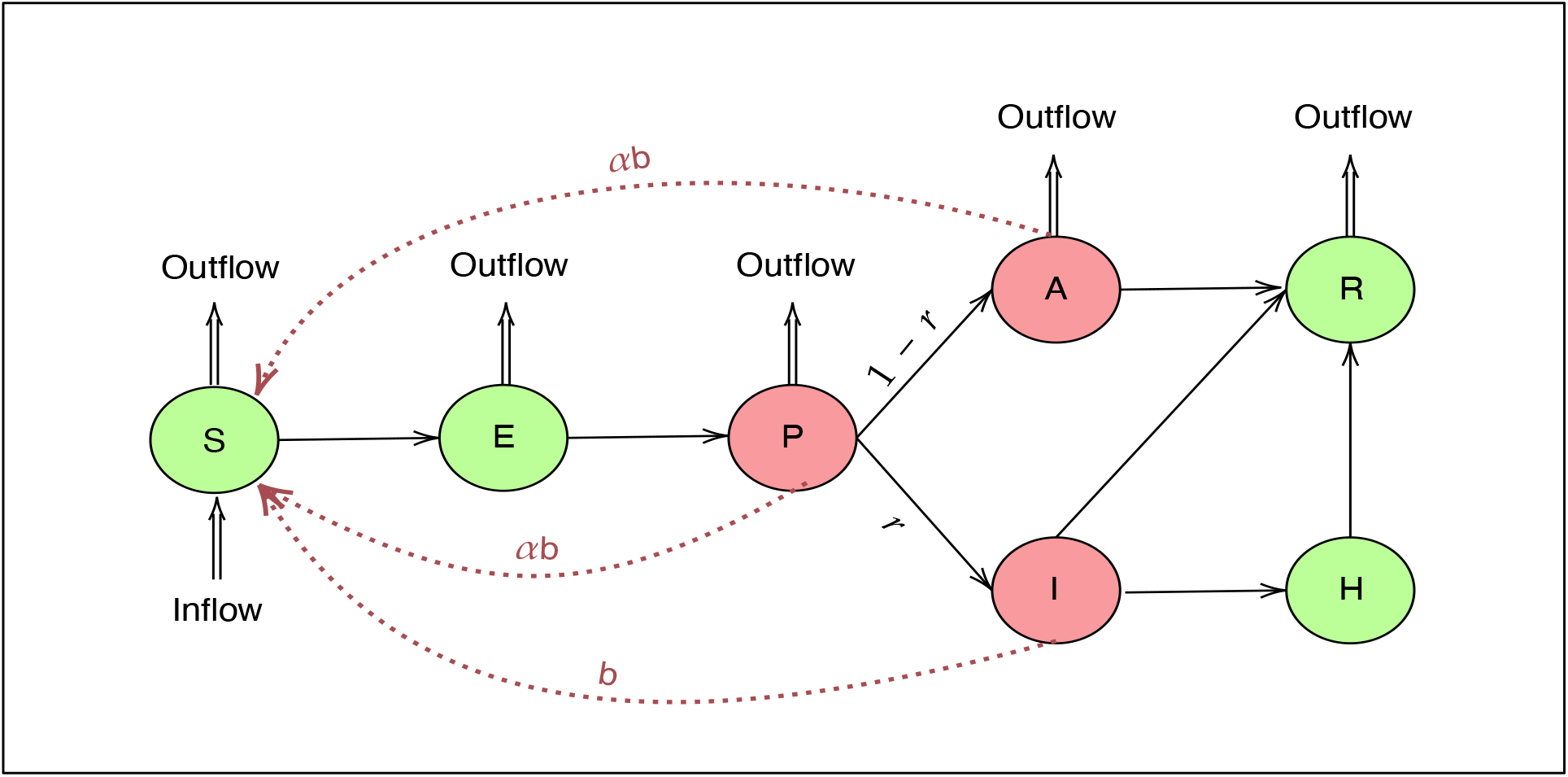
The SAPHIRE model with separate compartments for the latent, unascertained and ascertained cases.

##### Formulation

The dynamics of the 7 compartments described above at time *t* are described by the set of ordinary differential equations

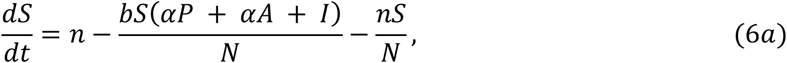

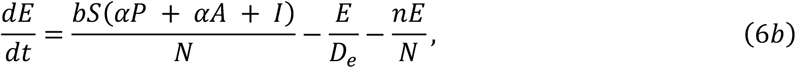

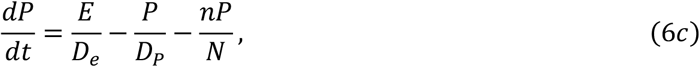

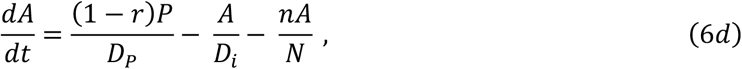

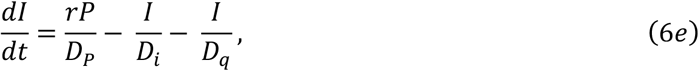

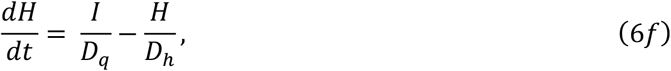

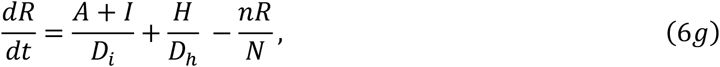

in which *b* is the transmission rate for ascertained cases (defined as the number of individuals that an ascertained case can infect per day), α is the ratio of the transmission rate of unascertained cases to that of ascertained cases, *r* is the ascertainment rate, *D*_*e*_ is the latent period, *D*_*p*_ is the pre-symptomatic infectious period, *D*_*i*_ is the symptomatic infectiousness period, *D*_*q*_ is the duration from illness onset to isolation and *D*_*h*_ is the isolation period in the hospital.

Under this setup, reproductive number *R* (as presented in the original manuscript) may be expressed as

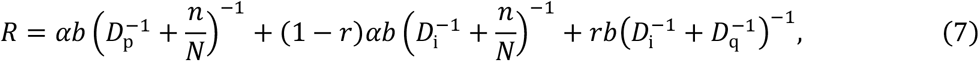

in which the three terms represent infections contributed by pre-symptomatic individuals, unascertained cases and ascertained cases, respectively. The model adjusts the infectious periods of each type of case by taking population movement (*n*/*N*) and isolation 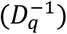 into account. For the case of India, we set *n* = 0.

##### Initial states and parameter settings

We set α = 0.55, assuming lower transmissibility for unascertained cases. Compartment *P* contains both ascertained and unascertained cases in the pre-symptomatic phase. We set the transmissibility of *P* to be the same as unascertained cases, because it has previously been reported that the majority of cases are unascertained (26). We assume an incubation period of 5.2 days and a pre-symptomatic infectious period *D*_*p*_ = 2.3 days. The latent period was *D*_*e*_ = 2.9 days. Because pre-symptomatic infectiousness was estimated to account for 44% of the total infections from ascertained cases (27), we set the mean of total infectious period as (*D*_*p*_ + *D*_*i*_) = *D*_*p*_/0.44 =5.2 days, assuming constant infectiousness across the pre-symptomatic and symptomatic phases of ascertained cases – thus the mean symptomatic infectious period was *D*_*i*_ = 2.9 days. We set a long isolation period of *D*_*h*_ = 30 days, but this parameter has no effect on the model fitting procedure, or the final parameter estimates. The duration from the onset of symptoms to isolation was estimated to be *D*_*q*_ = 7 as the median time length from onset to confirmed diagnosis in periods 1 - 4 respectively. On the basis of the parameter settings above, the initial state of the model is specified on March 15. The initial number of ascertained symptomatic cases *I*(0) is specified as the number ascertained cases in which individuals experienced symptom onset during 12-14 March. The initial ascertainment rate is assumed to be *r*_0_ (*r*_0_ = 2/100), and thus the initial number of unascertained cases is 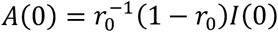. *P*_1_(0) and *E*_1_(0) denote the numbers of ascertained cases in which individuals experienced symptom onset during 15–16 March and 17–19 March, respectively. Then, the initial numbers of exposed and pre-symptomatic individuals are set as 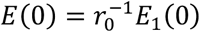 and 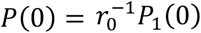, respectively. The initial number of the hospitalized cases *H*(0) is set as half of the cumulative ascertained cases on 8 March since *D*_*q*_ = 7 and there would be more severe cases among the ascertained cases in the early phase of the epidemic.

##### Likelihood and MCMC algorithm

Considering the time-varying strength of control measures implemented in India over the four lockdown periods, the model assumes that the value of *b* (*r*) corresponding to the *b*(*r*)lockdown period is *b*_*i*_ (*r*_*i*_) fo*r i* = 1,2,3, 4. The observed number of ascertained cases in which individuals experience symptom onset on day *t* – denoted by ∼_*R*_ – is assumed to follow a Poisson distribution with rate 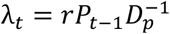, with *P*_t_ denoting the expected number of pre-symptomatic individuals on day *t*. The following likelihood equation is used to fit the model using observed data from March 15 (*T*_0_) to June 18 (*T*_1_).

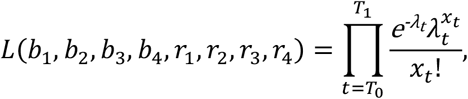

and the model is used to predict COVID-counts from June 19 to July 18. A non-informative prior of *U*(0,2) is used for *b*_1_, *b*_2_, *b*_3_ and *b*_4._. For *r*_1_, an informative prior of Beta(7.3, 24.6), is used, by matching the first two moments of the estimate using data from Singapore, as done by the authors of the SAPHIRE model. Re-parameterizing *r*_2_, *r*_3_ and *r*_4_ as

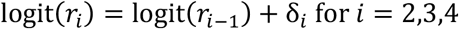

where logit(*t*) = log(*t*/(1 − *t*)) is the standard logit function. In the MCMC, δ_*i*_ ∼ *N*(0,1) for *i* = 2, 3, 4. A burn-in period of 100,000 iterations is fixed, with a total of 200,000 iterations being run.

#### 2.1.d. SEIR-fansy model

##### Overview

One of the problems with applying a standard SIR model in context of the COVID-19 pandemic is the presence of a long incubation period. As a result, extensions of SIR model like the SEIR model are more applicable. In the previous subsection we have seen an extension which includes the ‘asymptomatic infectious’ compartment (people who are infected and contributing to the spread of the virus, but do not show any symptoms). In this model, we use an alternate formulation by defining an ‘untested infectious’ compartment for infected people who are spreading infection but are not tested after the incubation period. This is necessary because there is a large proportion of infected people who are not being tested. We have assumed that after the ‘exposed’ node, a person enters the either ‘untested infectious’ node or the ‘tested infectious’ node. The ‘untested’ node mainly consists of the asymptomatic people. To incorporate the possible effect of misclassifications due to imperfect testing, we include a compartment for false negatives (infected people who are tested but reported as negative). As a result, after being tested, an infected person enters either into the ‘false negative’ node or the ‘tested positive’ node (infected people who are tested and reported to be positive). We keep separate nodes for the recovered and deceased persons coming from the untested and false negatives nodes which are ‘recovered unreported’ and ‘deceased unreported’ respectively. For the ‘tested positive’ node, the recovered and death nodes are denoted by ‘recovered reported’ and ‘deceased reported’ respectively. Thus, we divide the entire population into 10 main compartments: S (Susceptible), E (Exposed), T (Tested), U (Untested), P (Tested positive), F (Tested False Negative), RR (Reported Recovered), RU (Unreported Recovered), DR (Reported Deaths) and DU (Unreported Deaths).

##### Formulation

Like most compartmental models, this model assumes exponential times for the duration of an individual staying in a compartment. For simplicity, we approximate this continuous-time process by a discrete-time modeling process. The main parameters of this model are *β* (rate of transmission of infection by false negative individuals), *α*_*p*_ (ratio of rate of spread of infection by tested positive patients to that by false negatives), *α*_*H*_ (scaling factor for the rate of spread of infection by untested individuals), *D*_*e*_ (incubation period in days), *D*_*r*_ (mean days till recovery for positive individuals), *D*_*R*_ (mean number of days for the test result to come after a person is being tested), *μ*_c_ (death rate due to COVID-19 which is the inverse of the average number of days for death due to COVID-19 starting from the onset of disease times the probability death of an infected individual due to COVID), *λ* and *μ* (natural birth and death rates respectively, assumed to be equal for the sake of simplicity), *r* (probability of being tested for infectious individuals), *f* (false negative probability of RT-PCR test), *β*_1_ a*n*d 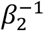 (scaling factors for rate of recovery for undetected and false negative individuals respectively), *β*_1_ *and* 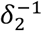 (scaling factors for death rate for undetected and false negative individuals respectively). The number of individuals at the time point *t* in each node is governed by the system of differential equations given by Equations (8a) – (8i). To simplify this model, we assume that testing is instantaneous. In other words, we assume there is no time difference from the onset of the disease after the incubation period to getting test results. This is a reasonable assumption to make as the time for testing is about 1-2 days which is much less than the mean duration of stay for the other compartments (further, once the person shows symptoms for COVID-19 like diseases, they are sent to get tested almost immediately). *Figure 4* provides a schematic overview of the model.

**Figure 4:**
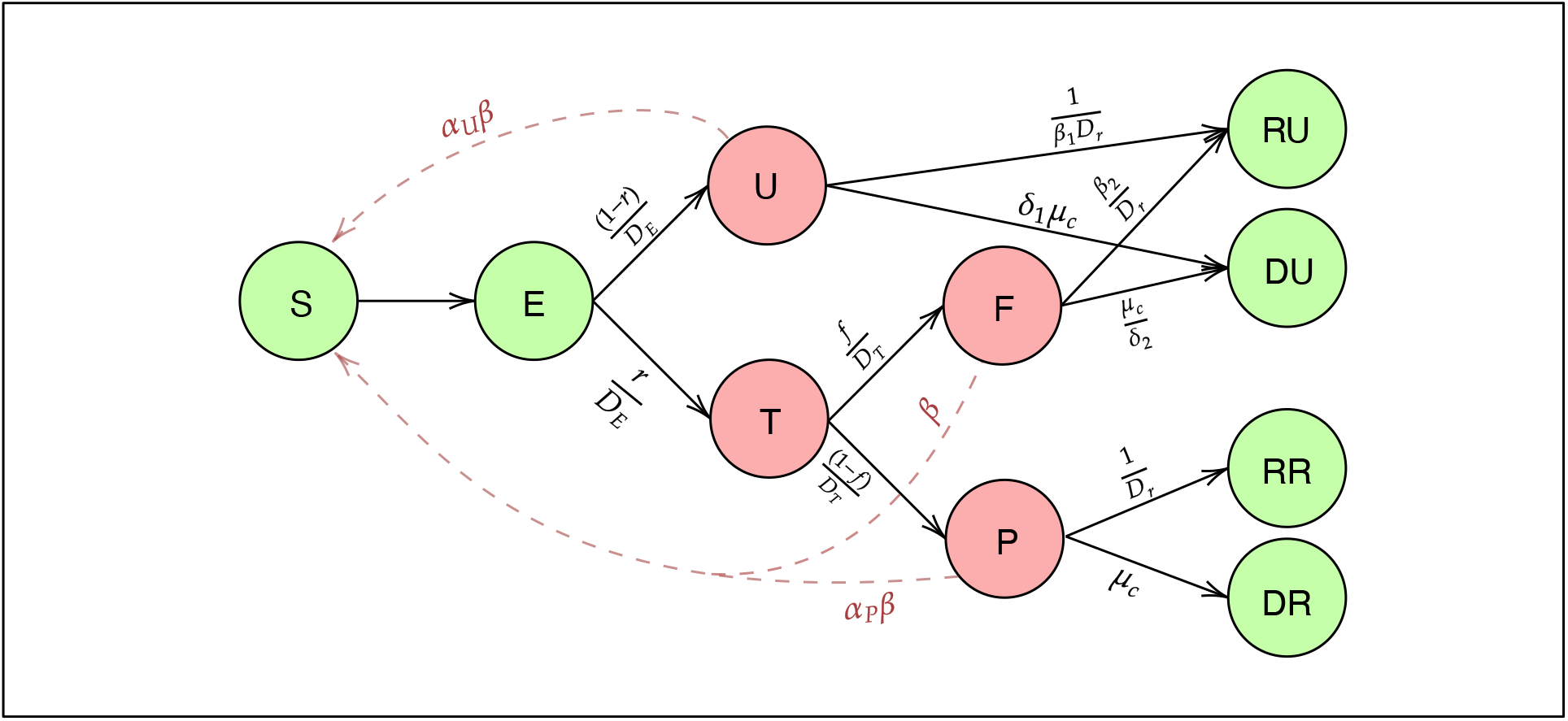
Schematic diagram for the SEIR-fansy model with imperfect testing and misclassification.

The following differential equations summarize the transmission dynamics being modeled.

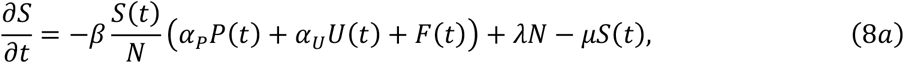

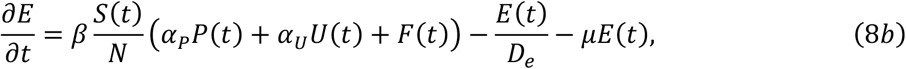

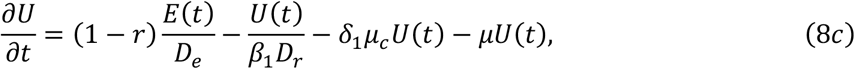

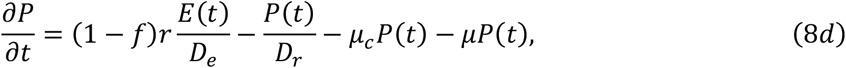

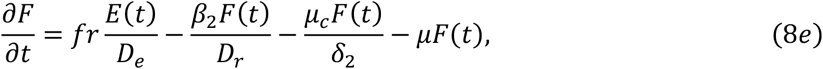

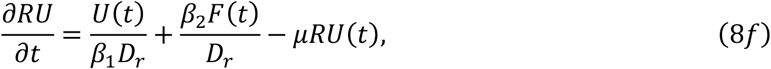

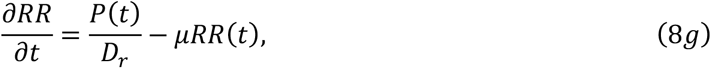

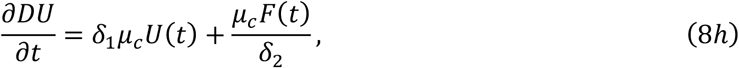

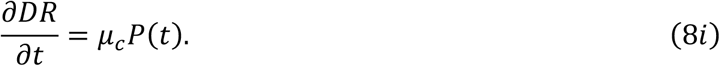

Using the Next Generation Matrix Method (28), we calculate the basic reproduction number

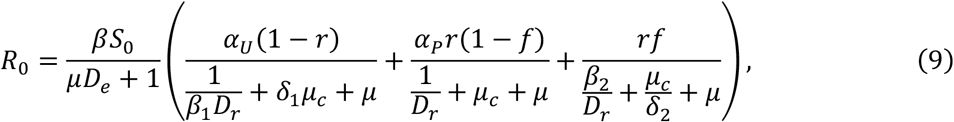

where *S*_0_ = λ/µ = 1 since we assume that natural birth and death rates are equal within this short period of time. *Supplementary Table S1* describes the parameters in greater detail.

##### Likelihood assumptions and estimation

Using Bayesian estimation techniques and MCMC methods (namely, Metropolis-Hastings method (29) with Gaussian proposal distribution) for estimating the parameters. First, we approximated the above set of differential equations using a discrete time approximation using daily differences. So, after we started with an initial value for each of the compartments on the day 1, using the discrete time recurrence relations we can find the counts for each of the compartments at the next days. To proceed with the MCMC-based estimation, we specify the likelihood explicitly. We assume (conditional on the parameters) the number of new confirmed cases on day *t* depend only on the number of exposed individuals on the previous day. Specifically, we use multinomial modeling to incorporate the data on recovered and deceased cases as well. The joint conditional distribution is

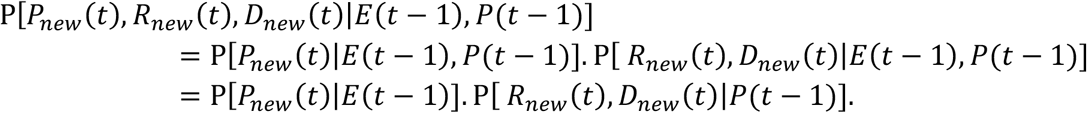

A multinomial distribution-like structure is then defined

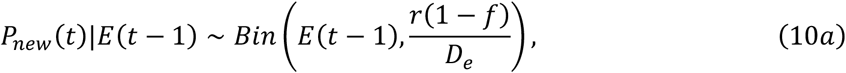

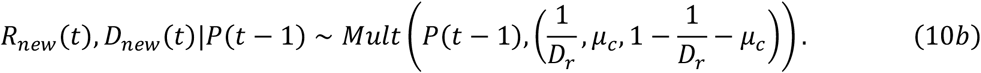

*Note:* the expected values of *E*(*t* − 1) and *P*(*t* − 1) are obtained by solving the discrete time differential equations specified by Equations (8a) – (8i).

##### Prior assumptions and MCMC

For the parameter *r*, we assume a *U*(0,1) prior, while for *β*, we assume an improper non-informative flat prior with the set of positive real numbers as support. After specifying the likelihood and the prior distributions of the parameters, we draw samples from the posterior distribution of the parameters using the Metropolis-Hastings algorithm with a Gaussian proposal distribution. We run the algorithm for 200,000 iterations with a burn-in period of 100,000. Finally, the mean of the parameters in each of the iterations are obtained as the final estimates of *β* and *r* for the different time periods.

#### 2.1.e. Imperial College London model (ICM)

##### Overview

Flaxman et al., 2020 introduce a Bayesian semi-mechanistic model for estimating the transmission intensity of SARS-CoV-2 (7). The model defines a renewal equation using the time-varying reproduction number *R*_*t*_ to generate new infections. As a lot of cases in SARS-CoV-2 are asymptomatic and reported case data is unreliable especially in early part of the epidemic in India, the model relies on observed deaths data and calculates backwards to infer the true number of infections. The latent daily infections are modeled as the product of *R*_*t*_ with a discrete convolution of the previous infections, weighted using an infection-to-transmission distribution specific to SARS-CoV-2. We implement this Bayesian semi-mechanistic model in the context of COVID-19 data arising from India in order to estimate the reproduction number over time, along with plausible upper and lower bounds (95% Bayesian credible intervals) of the daily infections and the daily number of infectious people. We parametrize *R*_*t*_ with a fixed effect and a random effect for each week over the course of the epidemic for each state. The fixed effect accounts for the variations in *R*_*t*_ across India as a whole whereas the random effect allows for variations among different states. The weekly effects are encoded as a random walk, where at each successive step the random effect has an equal chance of moving upwards or downwards from its current value. The model is implemented using epidemia (30), a general purpose R package for semi-mechanistic Bayesian modelling of epidemics. *Figure 5* represents a schematic overview of the model.

**Figure 5:**
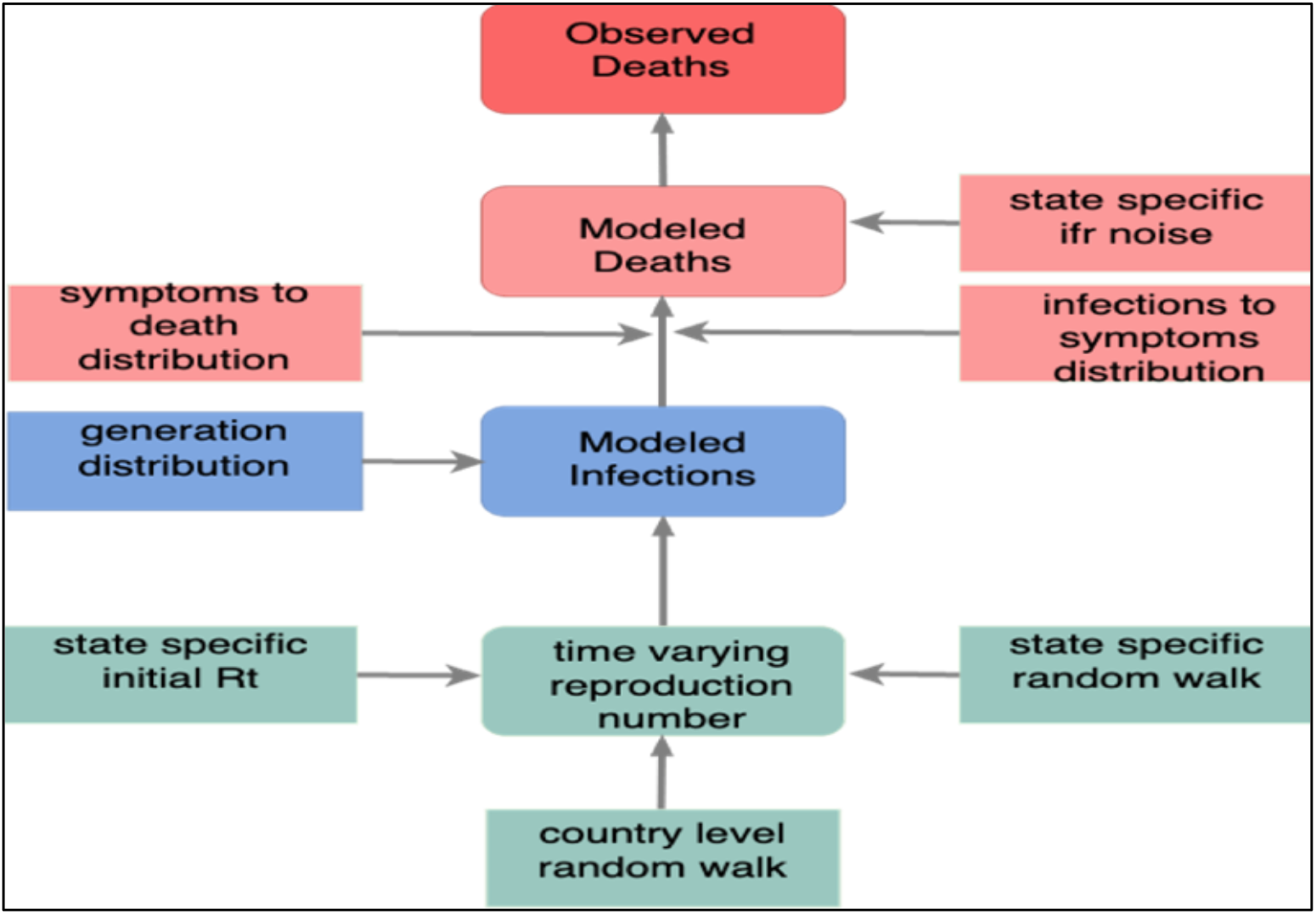
Schematic overview of ICM

##### Formulation

The true number of infected individuals, *i*, is modelled using a discrete renewal process. We specify a generation distribution (31) v with density *g*(τ) as *g* ∼ Gamma(6.5,0.62). Given the generation distribution, the number of infections *i*_*t,m*_ on a given day *t*, and state *m* is given by the following discrete convolution function:

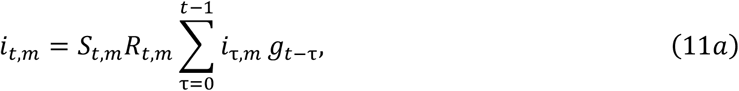

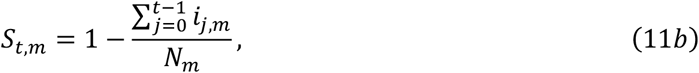

where the generation distribution is discretized by 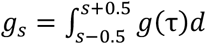 for *s* = 2,3, …, and 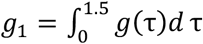. The population of state *m* is denoted by *N*_*m*_. We include the adjustment factor *S*_*t,m*_ to account for the number of susceptible individuals left in the population.

We define daily deaths, *D*_*t,m*_, for days *t* ∈ {1, …, *n*} and states *m* ∈ {1, …, *M*}. These daily deaths are modelled using a positive real-valued function *d*_*t,m*_ = *E*[*D*_*t,m*_] that represents the expected number of deaths attributed to COVID-19. The daily deaths *D*_*t,m*_ are assumed to follow a negative binomial distribution with mean *d*_*t,m*_ and variance 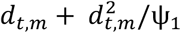, where ψ_1_ follows a positive half normal distribution, i.e.,

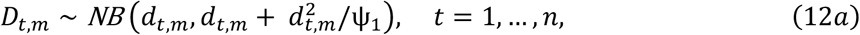

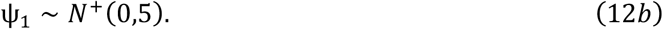

We link our observed deaths mechanistically to transmission(7). We use a previously estimated COVID-19 infection fatality ratio (IFR, probability of death given infection) of 0.01 together with a distribution of times from infection to death π. To incorporate the uncertainty inherent in this estimate we allow the ifr_m_ for every state to have additional noise around the mean. Specifically, we assume

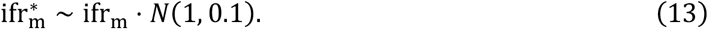

Using estimated epidemiological information from previous studies, we assume the distribution of times from infection to death π (infection-to-death) to be the convolution of an infection-to-onset distribution (π^′^) (32) and an onset-to-death distribution (24)

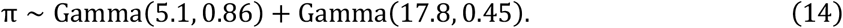

The expected number of deaths *d*_*t,m*_, on a given day *t*, for state *m* is given by the following discrete sum

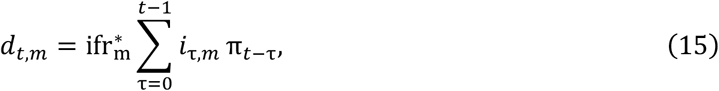

where *i*_τ,*m*_ is the number of new infections on day τ in state *m* and where, similar to the generation distribution, π is discretized via 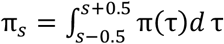 for *s* = 2,3, …, and 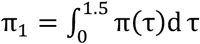, where π(τ) is the density of π.

We parametrize *R*_*t,m*_ with a random effect for each week of the epidemic as follows

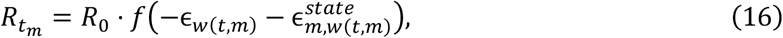

where *f*(*x*) = 2 *exp*(*x*) /(1 + *exp* (*x*)) is twice the inverse logit function, and ϵ_*w*(*t*)_ and 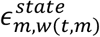 follows a weekly random walk (RW) process, that captures variation between *R*_*t,m*_ in each subsequent week. ϵ_*w*(*t*)_ is a fixed effect estimated across all the states and 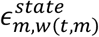 is the random effect specific to each state. The prior distribution for *R*_0_ (23) was chosen to be

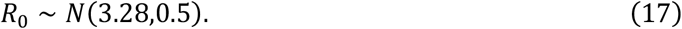

We assume that seeding of new infections begins 30 days before the day after a state has cumulatively observed 10 deaths. From this date, we seed our model with 6 sequential days of an equal number of infections: *i*_1_ = … = *i*_6_ ∼ Exponential(τ^−1^), where τ ∼ Exponential(0.03). These seed infections are inferred in our Bayesian posterior distribution.

We estimated parameters jointly for all 20 states. Fitting was done with the R package epidemia (30) which uses STAN (33), a probabilistic programming language, using an adaptive Hamiltonian Monte Carlo (HMC) sample.

### 2.2 Comparing models and evaluating performance

#### 2.2.a Choice of parameters

In order to implement the models described previously, we make certain assumptions (which are then utilized in back-calculations) in order to specify the model parameters. The initial reproduction number is fixed at *R* = 3.28(23). Unless mentioned otherwise, the mean time duration (in days) from infection to onset of symptoms is set at *D*_*e*_ = 5.1(34), while that from onset of symptoms to recovery is set at *D*_*r*_ = 17. 8 (13). The infection fatality rate was set at 0.01(24). The initial mean value of the removal rate is γ_0_ = 0.5436 and the proportion of death within the removed compartment is 0.01/0.5436 = 0.0184.

#### 2.2.b Metrics used for evaluation of models

Having established differences in the formulation of the different models, we compare their respective projections and inferences. In order to do so, we use the same data sources(35),(36) for all five models. Well-defined time points are used to denote training (March 15 to June 18) and test (June 19 to July 18) periods.

Using the parameter values specified above along with data from the training period as inputs, we compare the projections of the five models with observed data from the test period. In order to do so, we use the symmetric mean absolute prediction error (SMAPE) and mean squared relative prediction error (MSRPE) metrics as a measure of accuracy. Given observed time-varying data 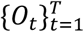 and an analogous time-series dataset of projections 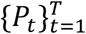, the SMAPE metric is defined as

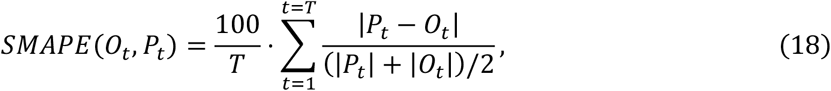

and the MSRPE is defined as

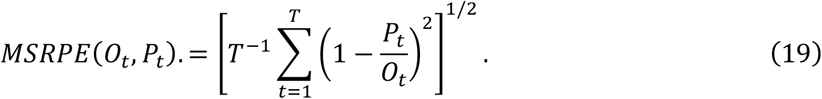

It can easily be seen that 0 ≤ *SMAPE* ≤ 100, with smaller values of both MSRPE and SMAPE indicating a more accurate fit. The baseline model yield projections of reported COVID-cases alone. The SAPHIRE and SEIR-*fansy* models return projections of *reported* and *unreported* COVID-cases and COVID-deaths *separately*. Finally, projections from ICM include true counts of COVID-cases (i.e., the sum of reported and unreported cases), in addition to true counts of COVID-deaths. *Supplementary Table S2* gives an overview of output from each of the models we consider.

In order to ensure a fair comparison, we compute the prediction error of *reported* projections from the models with respect to the observed data. Since the ICM projections are total counts (sum of reported and unreported), we do not include the same in this specific comparison method. For cumulative case counts, the model accuracies (SMAPE and MSRPE) are computed for all other models. For active cases (and cumulative deaths), accuracies of only the eSIR and SEIR-*fansy* models may be computed, as no other model returns projections for reported active case (and death) counts. Further, we compare (when possible) the estimated time-varying reproduction number *R*(*t*) over the four different stages of lockdown in India. Specifically, for each lockdown stage, we report the median *R*(*t*) value along with the associated 95% confidence interval CI. In addition to a systemic comparison of *R*(*t*), we also compare the projected active reported cases, cumulative cases and deaths on certain dates (specifically, June 30 and July 10) within the test period. The values are presented in *Table 2*.

**Table 1:** Overview of models studied.

| Name of model | Comments | Input(s) and output(s) | Parameter(s) and estimation |
| --- | --- | --- | --- |
| **Baseline**<br>(Bhardwaj, R. 2020) | Curve-fitting model.<br>Cumulative number of infected cases modeled as exponential process, with growth rate $\lambda$. | Daily time series of number of infected individuals from $T_0$ till $T_1^1$ (as input) and from $T_1$ to $T_2^2$ (as output). | Time varying growth rate of infection is estimated from input and modeled using least-squares regression. Maximum likelihood approach used for estimation. |
| **eSIR**<br>(Wang, L. et al., 2020) | Extension of the standard SIR<sup>2</sup> compartmental model. | Daily time series data on proportion of infected and recovered individuals from $T_0$ till $T_1^1$ (as input) and from $T_1$ to $T_2^2$ along with posterior distribution of parameters and prevalence values of the three compartments in the model (as output). | $\beta$ and $\gamma$ control transmission and removal rates respectively. $\lambda$ and $\kappa$ control variability of observed and latent processes respectively. Estimation involves implementing MCMC<sup>3</sup> methods for a hierarchical Bayesian framework. |
| **SAPHIRE**<br>(Hao, X. et al., 2020) | Extension of the standard SEIR<sup>2</sup> compartmental model. | Daily time series data from $T_0$ till $T_1^1$ on count of infected individuals (as input) and count of infected and removed individuals from $T_1$ to $T_2^2$ along with posterior distributions of parameters (as output). Unreported cases are also presented. | See Section 2.1.c for details on parameters. Estimation involves implementing MCMC<sup>3</sup> methods for a Bayesian framework. |
| **SEIR-fansy**<br>(Bhaduri, R., Kundu, R. et al., 2020) | Another extension of standard SEIR<sup>2</sup>, accounting for the possible effect of misclassifications due to imperfect testing. | Daily time series data from $T_0$ till $T_1^1$ on proportion of dead, infected and recovered individuals (as input) and from $T_1$ to $T_2^2$ along with posterior distributions of parameters and prevalence values of compartments in the model (as output). Unreported cases and deaths are also projected. | See **Supplementary Table S1** for details on parameters. Estimation involves implementing MCMC<sup>3</sup> methods for a hierarchical Bayesian framework. |
| **ICM**<br>(Flaxman et.al., 2020) | Renewal equation used to model infections as a latent process. Deaths are linked to infections via a survival distribution. Accounts for changes in behavior and various governmental policies enacted. | Daily time series data from $T_0$ till $T_1^1$ on count of dead individuals (as input) and from $T_1$ to $T_2$ (as output). Posterior over infections, deaths and various parameters. Infections include both symptomatic and asymptomatic ones. | See Section 2.1.e for details on parameters. Estimation is done via HMC<sup>4</sup> using STAN. |

(1) $T_0$ : time of crossing 50 confirmed cases – March 12, 2020. $T_1$ : June 18, 2020. $T_2$ : June 18, 2020.
(2) $S(E)IR$ : susceptible-(exposed)-infected-removed.
(3) MCMC: Markov chain-Monte Carlo.
(4) Hamiltonian Monte Carlo.

**Table 2:** Comparison of projections and prediction accuracies of the models under consideration.

|  |  | Model |  |  |  |  |
| --- | --- | --- | --- | --- | --- | --- |
|  |  | Baseline | eSIR | SAPHIRE | SEIR-fansy | ICM |
| Estimated mean reproduction number $R$ [95% CI] | **Lockdown 1.0**<br>*(March 25 – April 14)* | - | 2.08<br>[1.41, 2.12] | 2.08<br>[2.05, 2.11] | 4.09<br>[3.99, 4.20] | 1.41<br>[1.12, 1.77] |
|  | **Lockdown 2.0**<br>*(April 15 – May 3)* | - |  | 1.42<br>[1.40, 1.44] | 2.40<br>[2.37, 2.45] | 1.20<br>[0.92, 1.50] |
|  | **Lockdown 3.0**<br>*(May 4 – May 17)* | - |  | 1.24<br>[1.23, 1.26] | 1.78<br>[1.75, 1.83] | 1.29<br>[1.01, 1.59] |
|  | **Lockdown 4.0**<br>*(May 18 – May 31)* | - |  | 1.28<br>[1.27, 1.29] | 1.72<br>[1.70, 1.76] | 1.41<br>[1.11, 1.77] |
| Model-wise comparison of predicted and confirmed COVID-counts | **Active cases on June 30: 220,544**<br>*(on July 10: 284,212)* | - | 384,480<br>(811,430) | - | 218,688<br>(289,004) | - |
|  | **Cumulative cases on June 30: 585,481**<br>*(on July 10: 820,916)* | 568,564<br>(790,089) | 819,030<br>(1,649,776) | 568,074<br>(774,693) | 552,797<br>(762,742) | - |
|  | **Cumulative deaths on June 30: 17,411**<br>*(on July 10: 22,146)* | - | 25,596<br>(49,326) | - | 19,342<br>(27,432) | - |
| Model-wise comparison of prediction accuracy using SMAPE (MSRPE) | **Active cases** | - | 33.83<br>(1.55) | - | 0.72<br>(0.02) | - |
|  | **Cumulative cases** | 1.76<br>(0.03) | 23.10<br>(0.87) | 2.07<br>(0.05) | 3.20<br>(0.06) | - |
|  | **Cumulative deaths** | - | 26.30<br>(1.07) | - | 7.13<br>(0.19) | - |

Since we are interested in comparing relative performances of the models (specifically, their projections), we define another metric – the relative mean squared prediction error (Rel-MSPE). Given time series data on observed cumulative cases (or deaths) 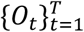, projections from a model A 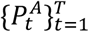, and projections from some other model B, 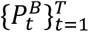, the Rel-MSPE of model B with respect to model A is defined as

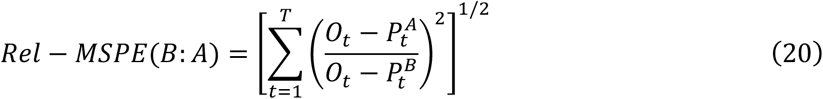

Since the baseline model yields projections of reported cumulative cases alone, we compute Rel-MSPE for the other models with respect to the baseline model for reported cumulative cases. Projections from ICM represent total (i.e., sum of reported and unreported) cumulative cases and deaths and are left out of this comparison of reported counts. For cumulative reported deaths, we compute Rel-MSPE of the SAPHIRE and SEIR-*fansy* and relative to the eSIR model. In addition to comparing the accuracy of fits that arise from the different models, we also investigate if projections from the different models are correlated with observed data. We use the standard Pearson’s correlation coefficient and Lin’s concordance correlation coefficient(37) as summary measures to study said correlation. Rel-MSPE and correlation metrics are presented in *Table 3*. As before, we carry out our comparisons based on the reported projections (and not the sum of reported and unreported projections) from the two SEIR models. No such consideration has to be made for the other three models.

**Table 3:** Comparison of relative performance and correlation with observed data of projections of the models under consideration.

| Observed data<br>(confirmed) | Metric | Model |  |  |  |  |
| --- | --- | --- | --- | --- | --- | --- |
|  |  | Baseline | eSIR | SAPHIRE | SEIR-*fansy* | ICM<sup>e</sup> |
| Cumulative cases | Rel-MSPE<sup>a</sup> | 1 | 0.225 | 3.819 | 0.579 | - |
|  | Pearson's correlation coefficient<sup>b</sup> | 1 | 0.985 | 1 | 1 | - |
|  | Lin's concordance coefficient<sup>b</sup> | 0.991 | 0.316 | 0.975 | 0.965 | - |
| Cumulative deaths | Rel-MSPE<sup>c</sup> | - | 1 | - | 7.605 | - |
|  | Pearson's correlation coefficient<sup>d</sup> | - | 0.978 | - | 0.999 | - |
|  | Lin's concordance coefficient<sup>d</sup> | - | 0.206 | - | 0.742 | - |

- a. For cumulative reported cases, Rel-MSPE is defined relative to projections from the baseline model. - b. For cumulative reported cases, the correlation coefficients of the projections are compared with respect to observed data. - c. For cumulative reported deaths, Rel-MSPE is defined relative to projections from the eSIR model. - d. For cumulative reported deaths, the correlation coefficients of the projections are compared with respect to observed data. - e. The ICM model returns total (reported + unreported) case and death counts, so we leave it out of our comparisons.

Since two models (SAPHIRE and SEIR-*fansy*) yield both reported as well as unreported counts of active or cumulative cases in addition to cumulative deaths, *Table 4* reports said counts on two specific dates – June 30 and July 10.

**Table 4:** Projected total (sum of reported and unreported) counts of cases (active and cumulative) and deaths (cumulative) from the SAPHIRE, SEIR-fansy and ICM models.

| Projected total count<sup>a</sup> | Projection from model on June 30<br>(and July 10) |  |  | Observed total count<sup>b</sup> | Underreporting factor<sup>c</sup> on June 30<br>(and July 10) |  |  |
| --- | --- | --- | --- | --- | --- | --- | --- |
|  | SAPHIRE | SEIR-fansy | ICM |  | SAPHIRE | SEIR-fansy | ICM |
| Active cases | - | 1,345,325<br>(1,773,330) | 798,204<br>(794,218) | 220,544<br>(284,212) | - | 6.10<br>(6.24) | 3.62<br>(2.79) |
| Cumulative cases | 16,270,126<br>(21,947,569) | 4,532,027<br>(6,178,763) | 5,354,425<br>(6,297,897) | 585,481<br>(820,916) | 27.79<br>(26.74) | 7.74<br>(7.53) | 9.15<br>(7.67) |
| Cumulative deaths | - | 63,112<br>(88,378) | 34,771<br>(44,231) | 17,411<br>(22,146) | - | 3.62<br>(3.99) | 2.00<br>(2.00) |

- a. Projected total count includes both reported as well as unreported values. - b. Observed total count represents only reported values. - c. Defined as projected total/observed reported counts, where total is the sum of reported and unreported cases.

### 2.3 Data source

The data on confirmed cases, recovered cases and deaths for India and the 20 states of interest are taken from COVID-19 India (35) and the JHU CSSE COVID-19 GitHub repository (36). In addition to this and other similar articles concerning the spread of this disease in India, we have created an interactive dashboard (38) summarizing COVID-19 data and forecasts for India and its states.

## 3. RESULTS

### 3.1. Estimation of reproduction number

From *Table 2*, we compare the mean of the time-varying effective reproduction number *R*(*t*) over the four phases of lockdown in India. The eSIR model does not return phase-specific values but returns a mean value of 2.08 (95% CI: 1.41 to 2.12) over the entire lockdown period. The mean (and 95% CI) values returned by the SAPHIRE model is 2.08 (95% CI: 2.05 to 2.11) during phase one of the lockdown, 1.42 (95% CI: 1.40 to 1.44) for phase two, 1.24 (95% CI: 1.23 to 1.26) for phase three and 1.28 (95% CI: 1.27 to 1.29) for the fourth and final lockdown phase. The SEIR-*fansy* notes that the mean drops from 4.09 (95% CI: 3.99 to 4.20) during the first phase of lockdown, to 1.72 (95% CI: 1.70 to 1.76) during the fourth lockdown phase. The ICM-based mean values fluctuate, from 1.41 (95% CI: 1.12 to 1.77) during the first lockdown phase, followed by 1.20 (95% CI: 0.92 to 1.50), then dropping to 1.29 (95% CI: 1.01 to 1.59) and finally rising to 1.41 again (95% CI: 1.11, 1.77) for the fourth phase of lockdown. In terms of agreement of reported values, SAPHIRE, SEIR-*fansy* and ICM report the highest mean *R* for phase one of the lockdown. While values reported by SEIR-*fansy* show a steady decrease over subsequent lockdown phases, both SAPHIRE and ICM report a drop in intermediate lockdown phases, followed by a rise. While SAPHIRE reports the lowest value of *R* for phase three, ICM reports the lowest value of *R* for phase two.

### 3.2 Estimation of reported case counts

From *Figure 6* and *Figure 9*, we note that the eSIR model overestimates the count of confirmed cumulative cases – a behavior which gets worse with time. The SAPHIRE, SEIR-*fansy* and baseline models slightly underestimate the count, with the baseline model performing the best, followed closely by SAPHIRE. This observation is supported by *Table 2* and *Table 3* as well – the projections from the baseline and SAPHIRE model on two specific dates (June 30 and July 10) are closer to the actual observed counts as compared to the other models. Additionally, the SMAPE and MSRPE values associated with the baseline model (1.76% and 0.03, respectively) are smaller than the other models. *Table 2* reveals a consistent behavior of model performance in terms of the SMAPE and MSRPE metrics, with the baseline model performing the best (SMAPE: 1.76%, MSRPE: 0.03), followed by the SAPHIRE model (SMAPE: 2.07%, MSRPE: 0.05), then the SEIR-*fansy* model (SMAPE: 3.20%, MSRPE: 0.06) and finally, the eSIR model (SMAPE: 23.10, MSRPE: 0.87). *Table 3* further reveals a similar comparison through Rel-MSPE values (all Rel-MSPE figures reported here are relative to the baseline model). The SAPHIRE model performs the best (Rel-MSPE: 3.819), followed by SEIR-*fansy* (with Rel-MSPE: 0.579), and finally, the eSIR model (Rel-MSPE: 0.225). All four sets of projections are highly correlated with the observed time series – with the baseline, SAPHIRE and SEIR-*fansy* models having a Pearson’s correlation coefficient of 1 with the observed data, while the eSIR model yields a value of 0.985. Lin’s concordance coefficient yields an ordering (from best to worst) of the baseline model (0.991), followed by the SAPHIRE model (0.975), the SEIR-fansy model (0.965) and finally, the eSIR model (0.316).

**Figure 6:**
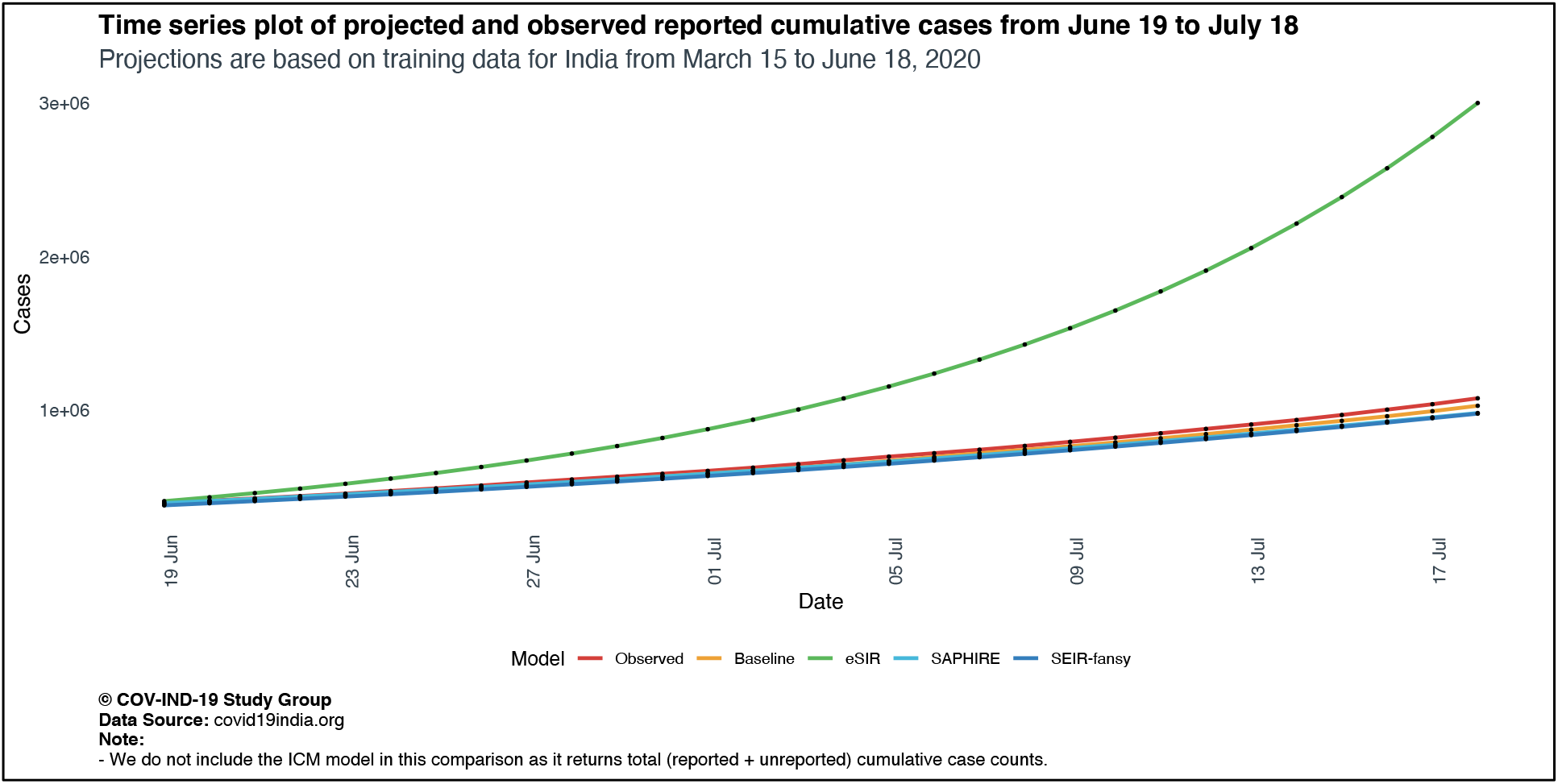
Comparison of projected and observed reported cumulative cases from June 19 to July 19 for India, using training data from March 15 to June 18.

Comparing confirmed active case counts across models, from *Figure 7* and *Figure 10*, we note a similar behavior, with the eSIR model consistently overestimating counts, while the SEIR-*fansy* model performs the best – *Table 2* and *Table 3* support this observation. The projection from the SEIR-*fansy* model are the closest to the actual observed values on June 30 and July 10. The eSIR model is much further off the mark. In terms of prediction accuracy, the SEIR-*fansy* model has an SMAPE value of 0.72% and an MSRPE value of 0.02. For eSIR model, those values are at 33.83% (SMAPE) and 1.55 (MSRPE).

**Figure 7:**
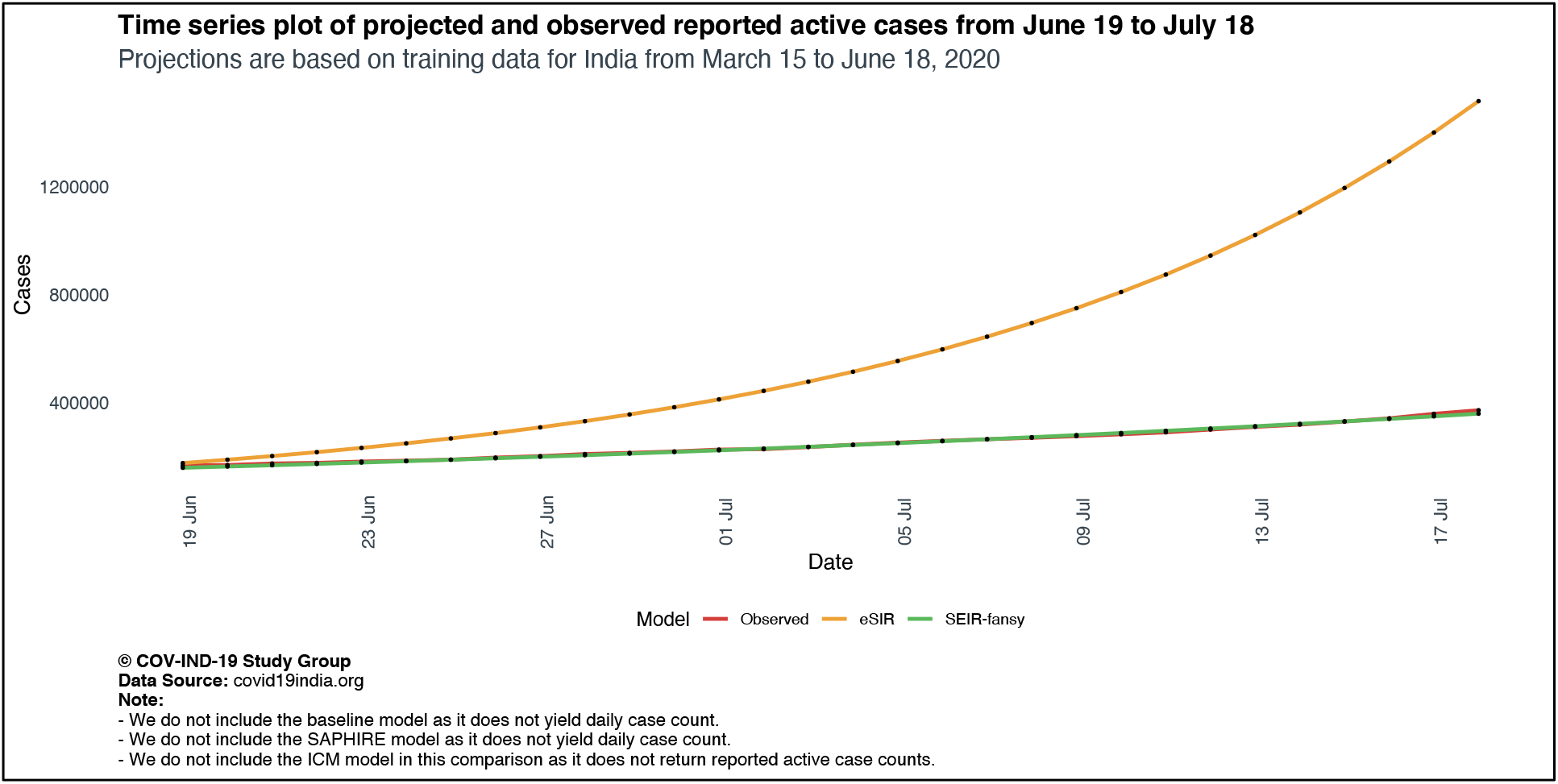
Comparison of projected and observed reported active cases from June 19 to July 19 for India, using training data from March 15 to June 18.

**Figure 8:**
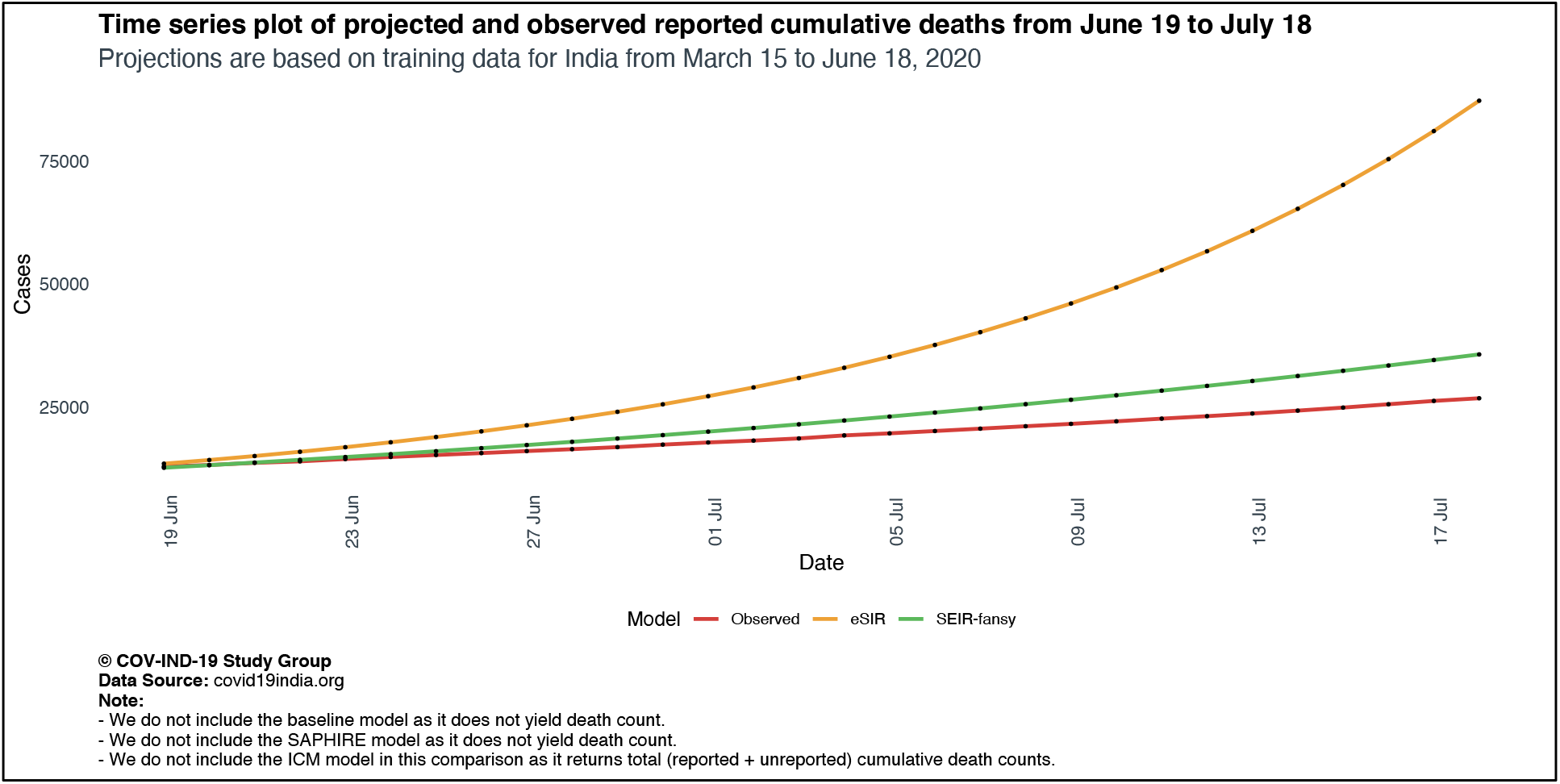
Comparison of projected and observed reported deaths from June 19 to July 19 for India, using training data from March 15 to June 18.

**Figure 9:**
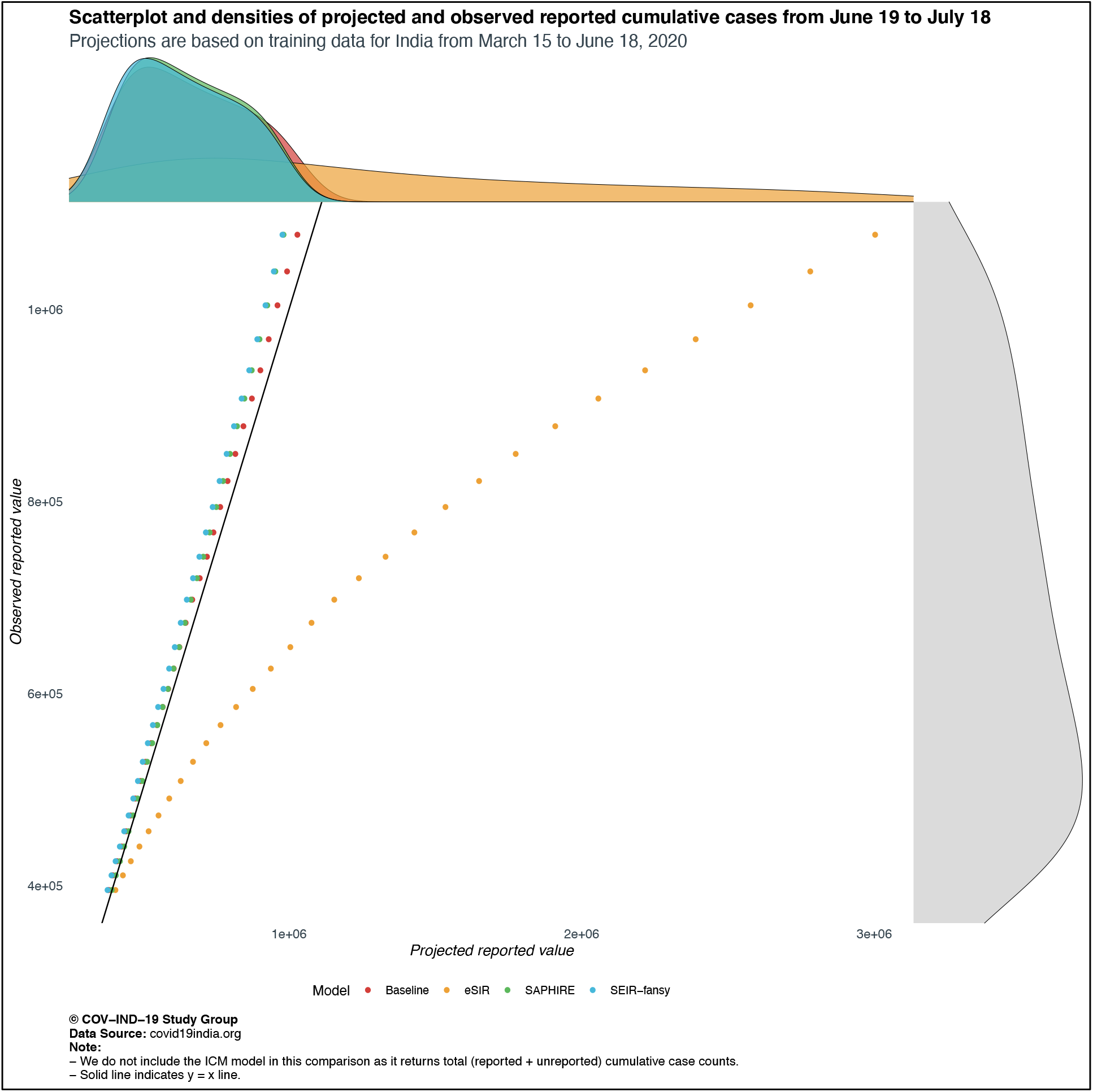
Scatter plot and marginal densities of projected and observed reported cumulative cases from June 19 to July 19 for India, using training data from March 15 to June 18.

**Figure 10:**
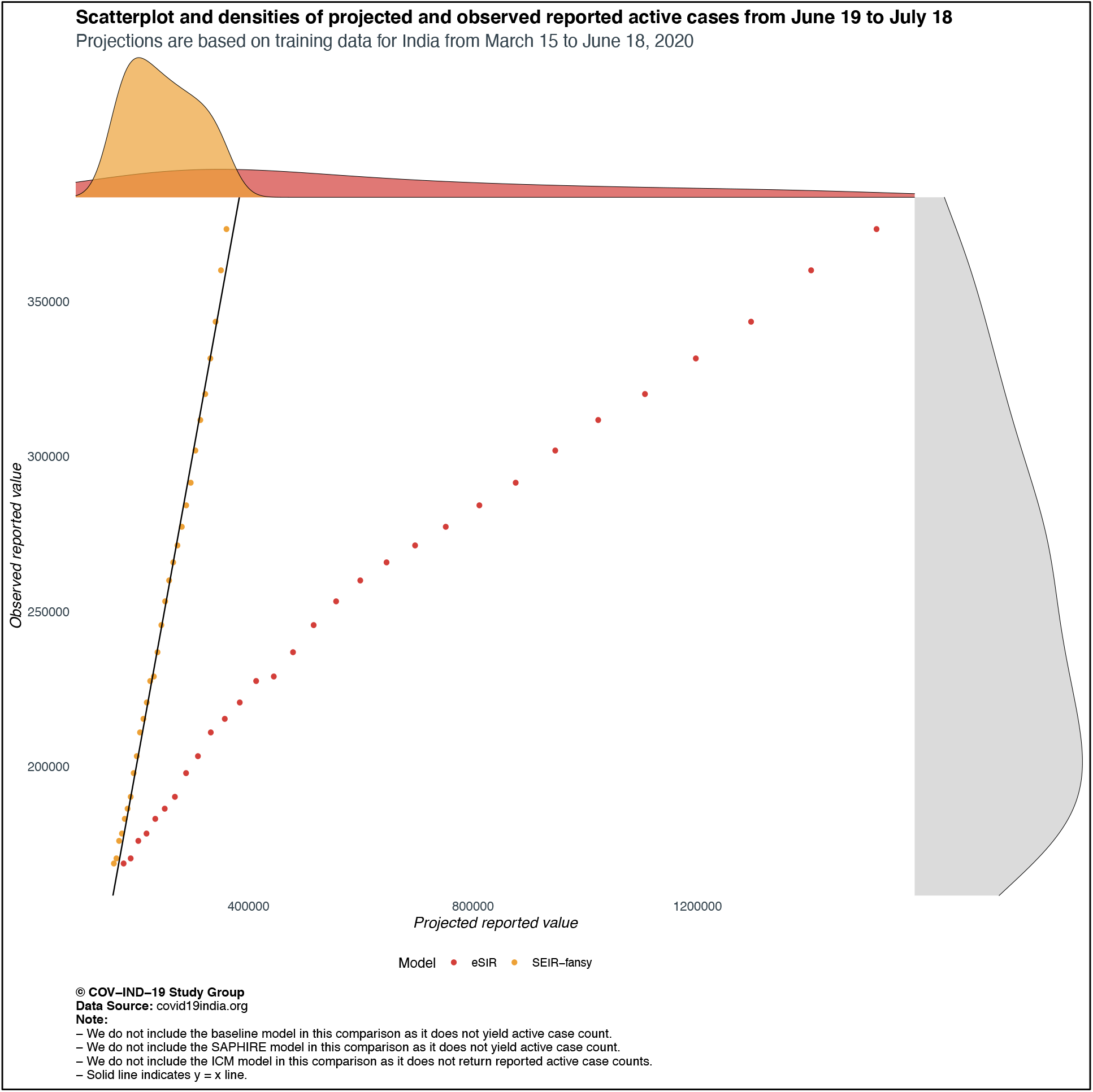
Scatter plot and marginal densities of projected and observed reported active cases from June 19 to July 19 for India, using training data from March 15 to June 18.

### 3.3. Estimation of reported death counts

From *Figure 8* and *Figure 11*, we note that the eSIR and SEIR-*fansy* models almost always overestimate the confirmed cumulative death counts. The eSIR model exhibits the poorest performance of the four models considered here – projecting an exponentially growing death count, whereas the observed data and projections from the SEIR-*fansy* model shows a linear-like trend. From *Table 2 and Table 3*, the SMAPE and MSRPE values, along with comparison of projections with observed data reveal that the SEIR-*fansy* model is the more accurate (SMAPE: 7.13%, MSRPE: 0.19) as compared to the eSIR model (SMAPE: 26.30%, MSRPE: 1.07). Relative to the eSIR model, the Rel-MSPE values of the models reveal that the SEIR-*fansy* model performs better (Rel-MSPE: 7.61). Judging by values of Pearson’s correlation coefficient, both sets of projections are highly correlated with the observed data. Lin’s concordance coefficient yields an ordering of SEIR-*fansy* (0.742), followed by eSIR (0.206).

**Figure 11:**
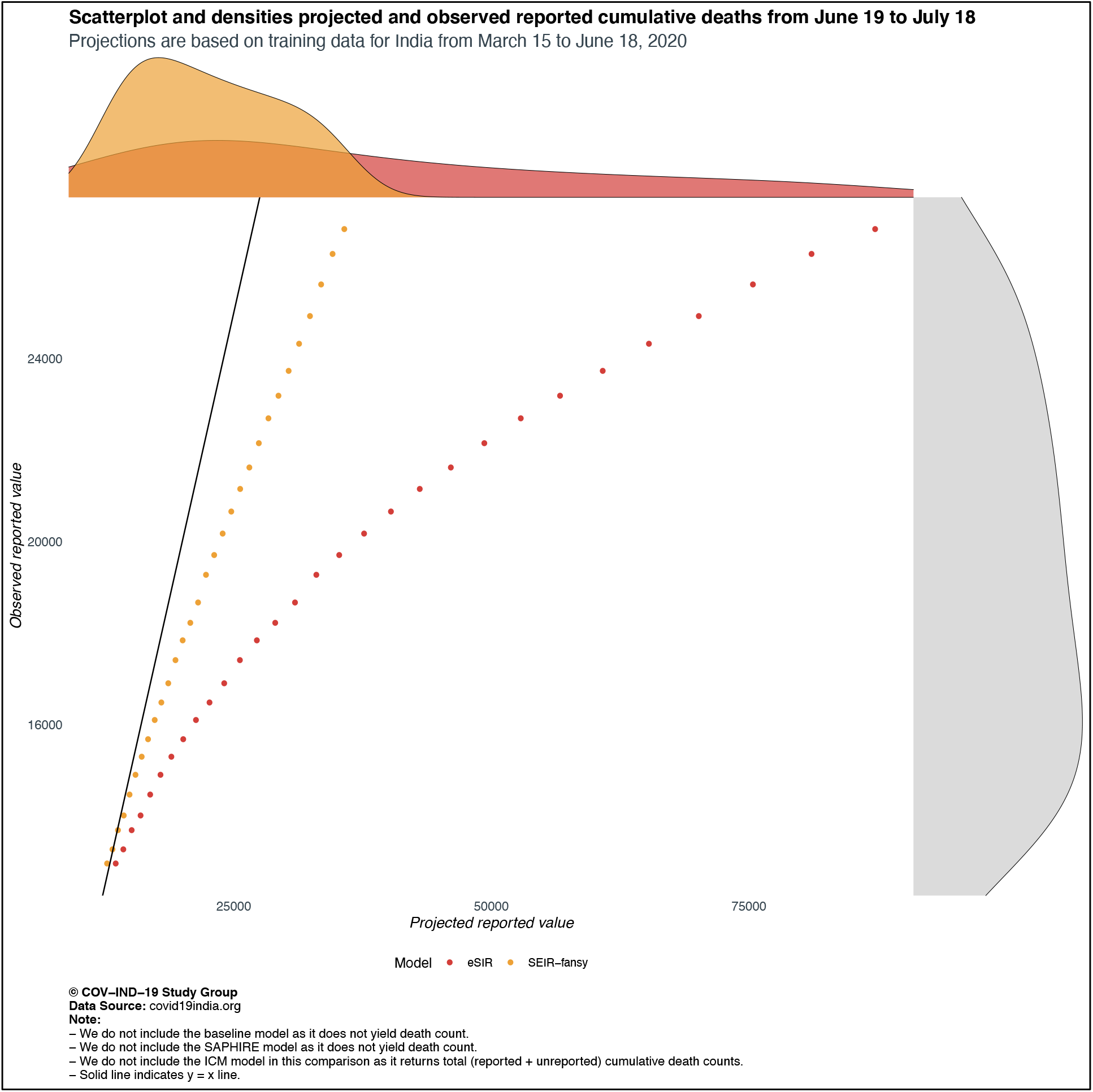
Scatter plot and marginal densities of projected and observed reported deaths from June 19 to July 19 for India, using training data from March 15 to June 18.

### 3.4. Estimation of unreported case and death counts

From *Table 4*, we observe that the SEIR-*fansy* model projects the maximum count of active cases and cumulative deaths on June 30 and July 10, followed by the ICM. The relative ordering of projections is reversed for cumulative cases, with the ICM projecting maximum case counts, followed by the SEIR-*fansy* model and finally, the SAPHIRE model. Comparing underreporting factors (total counts/observed counts), we note that the factors remain fairly comparable over time (June 30 vs July 10). For active case counts and cumulative death counts, the factor is higher for SEIR-*fansy* as compared to ICM. For cumulative case counts, SAPHIRE has the highest factor, followed by ICM and finally, SEIR-*fansy*.

## 4. DISCUSSION

In this comparative paper we have described five different models of various stochastic structures that have been used for modeling SARS-Cov-2 disease transmission in various countries across the world. We applied them to a case-study in modeling the full disease transmission of the coronavirus in India. While simulation studies are the only gold standard way to compare the accuracy of the models, here we were uniquely poised to compare the projected case-counts against observed data on a test period. We learned several things from these models. While the estimation of the reproduction number is relatively robust across the models, the prediction of daily active number of cases does show variation across models. The largest variability across models is observed in predicting the “total” number of infections including reported and unreported cases. The degree of underreporting has been a major concern in India and other countries(34). On two specific dates (June 30 and July 10), for cumulative case counts, we estimate the underreporting factors to be 27.79 and 26.74 respectively from the SAPHIRE model, 7.74 and 7.53 respectively from SEIR-*fansy* and 9.15 and 7.67 respectively from ICM. Similarly, for cumulative death counts, SEIR-*fansy* yields underreporting factors 3.62 on June 30 and 3.99 on July 10, while ICM notes that the underreporting factor is approximately 2.00 for both dates. With a comprehensive exposition and a single beta-testing case-study we hope this paper will be useful to understand the mathematical nuances and the differences in terms of deliverables for the models.

There are several limitations to this work. First and foremost, all model estimates are based on a scenario where we assumed no change in either interventions or behavior of people in the forecast period. This is not true as there is tremendous variation in policies across Indian states in the post lock-down phase. We did observe regional lockdowns that were enacted in the forecast period. None of our models tried to capture this variability. Second, the five models we compare are a subset of vast amount of work that has been done in this area, particularly models that incorporate age-specific contact network and spatiotemporal variation. Finally, an extensive simulation study would be the best way to assess the models under different scenarios but we have restricted our attention to India. Finally, we only report point estimates and have not compared the uncertainty estimates from each model which also play a key role in our choice.

## Data Availability

All data and code available at covind19.org

https://github.com/umich-cphds/cov-ind-19

## ACKNOWLEDGEMENTS

The authors would like to thank the Center for Precision Health Data Sciences at the University of Michigan School of Public Health, The University of Michigan Rogel Cancer Center and the Michigan Institute of Data Science for internal funding that supported this research. The authors are grateful to Professors Eric Fearon, Aubree Gordon and Parikshit Ghosh for useful conversations that helped formulating the ideas in this manuscript.

## DECLARATIONS

### Availability of data and material

Please visit https://github.com/umich-cphds/cov-ind-19

### Code availability

Please visit https://github.com/umich-cphds/cov-ind-19

### Authors’ contributions

Rupam Bhattacharyya and Maxwell Salvatore implemented the eSIR model. Xuelin Gu implemented the SAPHIRE model. Ritwik Bhadhuri, Ritoban Kundu and Bhramar Mukherjee developed the SEIR-*fansy* model. Ritwik Bhadhuri and Ritoban Kundu implemented the SEIR-*fansy* model. Swapnil Mishra was part of the group that developed ICM and implemented the same for this manuscript. Soumik Purkayastha implemented the baseline model. All authors participated in writing this manuscript. Bhramar Mukherjee conceptualized this project and oversaw the research.

---

**Supplementary Table S1:** Summary of initial values and parameter settings for application of the SEIR-fansy model in the context of COVID-19 data from India.

| Parameters | Settings | Description |
| --- | --- | --- |
| $\beta$ | Time-varying | Rate of infectious transmission by infected individuals with false negative test results. |
| $\alpha_p$ | 0.5 | Ratio of rate of spread of infection by patients who test positive, to rate of spread of infection by patients who get false negative results<sup>a</sup>. |
| $\alpha_U$ | 0.7 | Scaling factor for the rate of spread of infection by untested individuals<sup>a</sup>. |
| $D_e$ | 5.1 | Incubation period (in days). |
| $D_r$ | 17.8 | Recovery time (in days) for infected individuals. |
| $D_t$ | 0 | Waiting time (in days) for test result for tested individuals. |
| $\mu_c$ | 0.0562 | Death rate attributable to COVID-19<sup>b</sup>. |
| $\lambda, \mu$ | $3.95 \times 10^{-5}$ | Natural birth and death rates, respectively<sup>b</sup>. |
| $r$ | Time-varying | Probability of being tested for infectious individuals. |
| $f$ | 0.30 | Probability of a false negative RT-PCR diagnostic test result. |
| $\beta_1, \beta_2$ | 0.6 ($\beta_1$) and 0.7 ($\beta_2$) | Scaling factors for rate of recovery for undetected and false negative individuals respectively<sup>c</sup>. |
| $\delta_1, \delta_2$ | 0.3 ($\delta_1$) and 0.7 ($\delta_2$) | Scaling factors for death rate for undetected and false negative individuals respectively<sup>f</sup>. |

- $\alpha_p < 1$ represents the scenario where individuals who test positive are infecting susceptible individuals at a lower rate than infected individuals with false negative test results. $\alpha_U < 1$ is assumed as U mostly consists of asymptomatic or mildly symptomatic cases who are known to spread the disease at a much lower rate than those with higher levels of symptoms. - Equal to the inverse of the average number of days for death starting from the onset of disease, times the probability of death of an infected individual. Natural birth and death rates are assumed to be equal for simplicity. - $\beta_1 < 1$ , $\beta_2 < 1$ are assumed, since the recovery rate is slower for individuals with false negative test results as compared to those who have been hospitalized. The condition of untested individuals is not as severe as they consist of mostly asymptomatic people. Consequently, they are assumed to recover faster than those with positive test results.
- d. $\delta_1 < 1$ , $\delta_2 < 1$ are assumed. The death rate for those with false negative test results is assumed to be higher than those with positive test results, since the former are not receiving proper treatment. For untested individuals, the death rate is taken to be lesser because they are mostly asymptomatic. As a result, their survival probability is much higher.

**Supplementary Table S2:** Overview of projected COVID-counts for each model considered.

| Type of count projected | COVID-counts |  |  |
| --- | --- | --- | --- |
|  | Cumulative<br>COVID-cases | Active<br>COVID-cases | Cumulative<br>COVID-deaths |
| **Reported** | Baseline, eSIR, SAPHIRE, SEIR-*fansy* | eSIR, SEIR-*fansy* | eSIR, SEIR-*fansy* |
| **Unreported** | SAPHIRE, SEIR-*fansy* | SEIR-*fansy* | SEIR-*fansy* |
| **Total**<br>**(reported + unreported)** | SAPHIRE, SEIR-*fansy*, ICM | SAPHIRE, SEIR-*fansy*, ICM | SEIR-*fansy*, ICM |

